# Rx Norm for Europe - Toward the representation of medicinal products in the OMOP CDM: Graph visualization and validation of two mapping approaches using the OHDSI USAGI tool and LLM

**DOI:** 10.64898/2026.01.15.26344216

**Authors:** Karen Triep, Marcel Messerli, Olga Endrich

## Abstract

Medication product names in Swiss electronic health records are heterogeneous and often encode multiple attributes (e.g., ingredient, strength, dose form, packaging) in German free text. This limits interoperability and reduces the utility of ATC codes, which do not uniquely identify products. We compared two workflows for mapping Swiss medication products to RxNorm and RxNorm Extension: (i) an Observational Health Data Sciences and Informatics (OHDSI) USAGI workflow with lexical similarity and expert curation, and (ii) a large language model (LLM) workflow with constrained synonym generation and candidate selection. The LLM workflow applied explicit attribute priorities and allowed abstention when no suitable match was available. Mapping results were loaded into a Neo4j graph database. We assessed semantic proximity using the median shortest path length between mapped concepts. We evaluated 179 products; 151 products were not equally mappable at code level. For these discordant products, the LLM workflow mapped predominantly to branded-level classes (121/151, 80.1 percent), whereas manual/USAGI mapping more often selected clinical drug–level classes (87/151, 57.6 percent). Semantic proximity differed by target vocabulary. In RxNorm, the LLM workflow achieved a lower overall median path length than manual/USAGI (2.47 vs 2.81), in RxNorm Extension, manual/USAGI achieved a lower median path length than LLM (2.46 vs 2.66). Graph-based inspection supported identification of ambiguous cases and systematic differences in hierarchical level. The results show that LLM-assisted mapping can be efficient and competitive; performance depends on the target vocabulary and concept class. Improved European coverage in RxNorm extensions remains necessary for standardization.

## 1. Introduction

Regional medication product naming has often evolved historically, resulting in heterogeneous structures, inconsistent naming, and reliance on national languages. At the product level, these names frequently conflate information from multiple domains—such as active ingredient(s), strength, dose form, pack size, route of administration, manufacturer, and occasionally even indications or marketing descriptors—making a clean one-to-one representation in interoperable terminologies challenging. This heterogeneity becomes particularly problematic when products must be represented consistently across institutions and languages, and when source strings need to be translated and normalized into English to be processed effectively by standard, term-based mapping tools.[1] Although ATC codes (Anatomical Therapeutic Chemical Classification System, controlled by the World Health Organization Collaborating Centre for Drug Statistics Methodology WHOCC) are widely used and highly valuable for pharmacological classification, they were not designed to uniquely identify medication products at the level typically required for real-world data standardization.[2] [3] It divides drugs into different groups according to the organ or system on which they act, their therapeutic intent or nature, and the drug’s chemical characteristics. ATC captures therapeutic class and, in many cases, the active ingredient, but it does not reliably encode critical product-defining attributes. As a result, ATC-based mappings can be ambiguous: multiple clinically distinct products may share the same ATC code, and ATC does not consistently support precise linkage to branded or pack-level concepts. This limits its utility for harmonizing medication exposure data in observational research settings.

For these reasons, mapping to RxNorm (“Rx” = medical prescription symbol and “Norm” = normalized naming system) is often more useful for interoperability and analytics, because RxNorm provides normalized identifiers across ingredient, clinical drug, branded drug, and—in combination with extensions—often pack-level representations.[4] RxNorm is part of Unified Medical Language System (UMLS) terminology and is maintained by the United States National Library of Medicine (NLM). Access is provided by APIs. Importantly, RxNorm serves as a practical “hub vocabulary”: once a source product is linked to an RxNorm concept, a range of downstream crosswalks can already be leveraged through existing mappings in the OMOP vocabulary ecosystem and in the norm itself.[5] [1] In practice, an RxNorm code can act as a pivot to other commonly used drug terminologies and groupers (for example, ingredient-level representations and drug class groupings), enabling consistent integration into interoperable data models such as the Observational Medical Outcomes Partnership (OMOP) Common Data Model (CDM)[6][7] without repeatedly rebuilding bespoke cross-mappings from national product names.

However, RxNorm coverage is not complete for the European[8] —and specifically Swiss—market. Many locally marketed products, presentations, and naming conventions are absent or only partially represented, which creates gaps when attempting to standardize non-U.S. medication data using RxNorm alone. This motivates the need for a European extension to RxNorm that explicitly addresses products and presentations not available in the U.S.-centric source, while preserving compatibility with OMOP standard concept conventions. Such an extension would help bridge market- and regulatory-specific differences, reduce reliance on fragile string matching and ad hoc local coding, and improve reproducibility of cross-site medication analyses.

With data interoperability gaining importance in healthcare delivery and international research, robust mapping support is essential.[1] The Observational Health Data Sciences and Informatics (OHDSI) community provides USAGI to support semi-automated mapping of source vocabularies to OMOP CDM standard concepts.[9] USAGI is the OHDSI community’s semi-automated mapping tool for aligning local source vocabularies (e.g., hospital medication lists) to OMOP CDM concepts by combining string-based similarity, configurable normalization, and an interactive curation interface. Practically, USAGI takes a source term table and a target concept vocabulary (which can include OMOP CDM standard vocabularies)[5], computes similarity scores between source strings and candidate target concept names / synonyms, and presents ranked candidates for human review, acceptance, and refinement. The resulting output is typically an explicit mapping table that can be represented as a strict one-to-one link (one source record mapped to a single target concept) or as a one-to-many set of links when multiple target concepts are required to represent a single source product. Because OMOP CDM vocabularies are often hierarchical, USAGI-supported targets can span different levels of granularity—ingredient, clinical drug, branded drug, and, where available, pack representations—allowing users to choose the level that best matches the information present in the source string. In addition, mappings may be produced in parallel to different classification schemes, for example linking the same source product to RxNorm and RxNorm Extension for interoperability, to ATC for pharmacologic grouping, and to SNOMED CT (Systematized Nomenclature of Medicine Clinical Terms) [10] where clinical or substance concepts are needed for downstream phenotyping or semantic integration. This is particularly useful when a single downstream workflow needs both a product identifier for medication exposure and a higher-level grouping variable for cohort definitions or stratified analyses. Importantly, USAGI is not limited to “standard vocabularies” in the narrow sense: it can also be used to map to non-standard or bespoke terminologies, provided the target concepts are available in a compatible table format with stable identifiers and terms.. As a result, USAGI can function both as a bridge into OMOP CDM standard concepts and as a general-purpose curation environment for building and maintaining mappings.

In this study, local medication products were mapped to RxNorm and RxNorm Extension concepts using two complementary approaches: (1) a conventional OHDSI USAGI workflow based on lexical similarity with structured manual review, and (2) a Large Language Model (LLM)-based workflow that proposes and selects candidate standard concepts under explicit attribute-matching constraints. Both pipelines were applied to the same curated reference set of product entries and mapping accuracy was assessed against the reference standard. The LLM workflow was designed to prioritize correctness of active ingredient and strength, while only matching manufacturer and pack size when these attributes were explicitly present in the source string. In parallel, USAGI mappings followed established similarity scoring and reviewer adjudication to resolve ambiguous or underspecified strings. The resulting mappings were enriched with hierarchical and compositional relationships across Swiss product entries and RxNorm and RxNorm Extension concepts to preserve both product-level and ingredient-level structure. All mappings and relationships—annotated by whether they originated from the USAGI or LLM approach—were ingested into a Neo4j [11] graph database. Using Neo4j Bloom, we explored interactive graph views to inspect mapping provenance, identify clusters of ambiguous products, and surface systematic error modes (e.g., strength mismatches or over-specific pack-level assignments). This graph-based representation enables quality inspection beyond record-by-record review by highlighting inconsistencies and many-to-one or one-to-many mapping patterns across the dataset. Overall, the objective is to maximize mapping precision for ingredient and strength while minimizing erroneous matches driven by brand, manufacturer, or packaging descriptors, and to make residual ambiguity transparent and audit-able within an interoperable framework. Moreover, his representation enabled a knowledge graph for medication.

Aim and Objective: We address the task of mapping free-text medication names from an electronic health record (EHR) integrated product list —primarily in German—to standardized RxNorm or RxNorm Extension concepts. The objective is to maximize mapping precision with respect to active ingredient, strength, and, when present, manufacturer and package size.

## 2. Methods

In this study, Swiss medication product entries were mapped to RxNorm and RxNorm Extension concepts using two complementary approaches: (i) a conventional OHDSI USAGI–based semi-automated mapping workflow and (ii) a large language model (LLM)–assisted workflow that generates and selects candidate standard concepts under explicit, attribute-level matching constraints. USAGI [9] is an OHDSI-supported mapping environment that operationalizes lexical similarity scoring, configurable string normalization, and interactive expert review and editing to align (local) source vocabularies with OMOP concepts and standard terminologies and classifications..

### 2.0.1. Data sources and scope

The input records consist of free text medication names in German originating from the clinical system. Target concepts are the RxNorm (United States) and RxNorm Extension (international coverage) vocabularies. To focus the mapping effort on products with practical clinical relevance, products were prioritized based on observed administration frequency derived from local medication usage records. Products were ranked by frequency and a subset of effectively administered products was selected for mapping. This selection step reduced noise from rarely used or obsolete catalog entries and ensured that the mapping pipeline was evaluated primarily on products contributing meaningfully to routine prescribing workflows.

### 2.1. USAGI assisted workflow

#### 2.1.1. Preparation of input files (CSV)

Medication-related terms were extracted from a local EHR product master and exported as a comma-separated values (CSV) input file. The export preserved the original product list fields required for downstream processing. All values were retained in their native representation to avoid introducing transformation bias prior to mapping. Basic normalization was applied only to support automated processing (e.g., trimming whitespace, harmonizing decimal separators, and standardizing unit spelling), while the original raw fields were kept unchanged in the output for traceability.

#### 2.1.2. Automated mapping using a usage-term similarity approach

Automated mapping from local EHR product terms to standardized medication concepts was performed using the term similarity approach against RxNorm and RxNorm Extension concept labels. For each local product entry, candidate RxNorm concepts were retrieved and scored based on lexical and structural similarity between the local usage term(s) and the candidate terminology strings. Candidate selection and ranking were guided with particular emphasis on extracting and comparing the ingredient and quantitative attributes when available.

#### 2.1.3. Closest-match definition and attribute prioritization

The “closest match” for each input product was determined using a hierarchical attribute priority rule to reflect clinically meaningful equivalence. Matches were evaluated in the following order of precedence: ingredient (highest priority), amount (e.g., strength as stated), concentration (e.g., mg/mL), and volume (e.g., total volume or package volume). Under this rule, candidate concepts sharing the same active ingredient were preferred over candidates that matched only quantitative or packaging attributes. Quantitative agreement was used to differentiate among candidates with the same ingredient, and volume served as a lower-priority tie-breaker.

#### 2.1.4. Output file structure and mapping quality rating

The mapping results were written to an output CSV containing, for each input record: the original input data fields (unchanged), the selected RxNorm and/or RxNorm Extension identifier(s), and a mapping quality rating representing the relationship between the local term and the selected standard concept. The rating categories were: equal (exact semantic match), equivalent (minor representational differences without semantic loss), narrower (selected concept more specific than the input), wider (selected concept more general than the input), and no match (no acceptable candidate identified). This structure ensured end-to-end auditing.

### 2.2. LLM assisted workflow

The pipeline takes German free-text medication strings from clinical systems and does not use proprietary brand names as matching keys. This improves robustness and reduces reliance on local branding artifacts that are unstable or not internationally interpretable. We implement a two-stage LLM-assisted pipeline followed by deterministic validation, separating recall-oriented retrieval from precision-oriented selection. In stage 1, an LLM generates up to three English synonyms per input, preserving active ingredient and strength when explicitly present while removing manufacturer and packaging details. The model is constrained to avoid hallucination: ambiguous strength or pack size is omitted rather than inferred, and strengths are never altered. These English synonyms increase lexical compatibility with RxNorm indexing and standard full-text search heuristics. In stage 2, we query RxNorm and the RxNorm Extension using the original German string plus up to three English synonyms, retrieving the top 20 candidates per query. Candidates are merged and deduplicated by concept identifier to form a consolidated candidate list per record. A second LLM then performs constrained re-ranking over this fixed list and outputs only a single index or a designated “no match” value. Selection prioritizes ingredient match and enforces strength consistency when present, deprioritizing candidates that introduce unsupported manufacturer or package constraints, with deterministic post-checks flagging out-of-range outputs for review see Figure 1.

**Figure 1.**
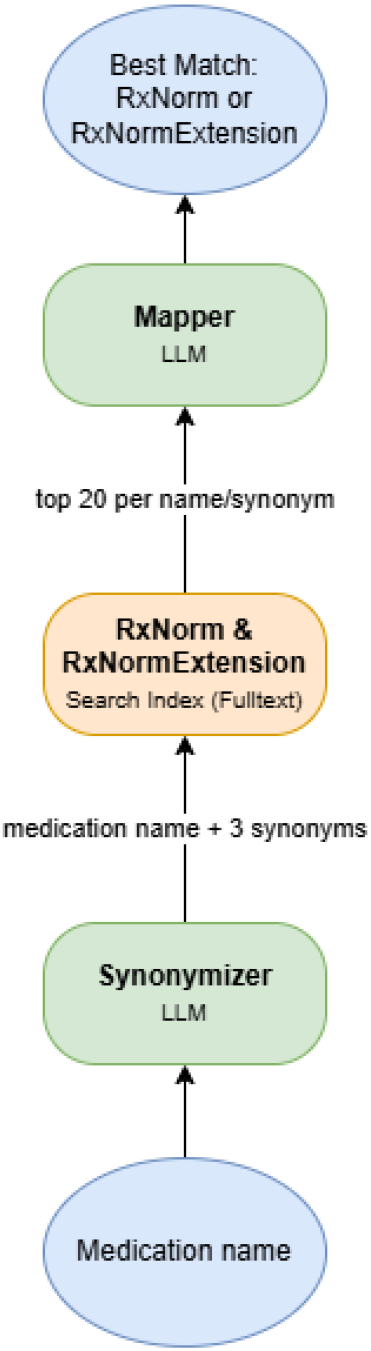
Workflow of the LLM approach

#### 2.2.1. Overall approach

All processing occurs without using proprietary brand names as matching keys We implement a two stage LLM assisted pipeline using GPT-4.1 via Microsoft Azure / OpenAI followed by a deterministic selection step:

##### 1. LLM assisted synonym generation (German → English)

- Goal. Create up to three English synonyms per input drug name that (a) preserve the active ingredient and, if present, the dose, but (b) remove manufacturer and packaging details; if dose/pack size information is ambiguous, omit it rather than hallucinating or altering it.
- Rationale. English synonyms increase recall and lexical compatibility with RxNorm/RxNorm Extension indexing and with full text search heuristics.
- System prompt constraints. The LLM is instructed to:

- Translate or paraphrase German medication names into English synonyms.
- Exclude brand names and manufacturer strings from the synonyms.
- Preserve strength if clearly present; otherwise omit it (never change a strength).
- Omit package size unless it is clearly required and consistent; do not invent or alter quantities.
- Return up to three concise synonyms per input.

##### 2. Full text retrieval over RxNorm / RxNorm Extension

- For each input name and each of its up to three English synonyms, we run a full text search against RxNorm/RxNorm Extension to retrieve the top 20 candidates per query (deduplicated across queries).
- We keep the candidate’s concept identifier

##### 3. LLM based candidate selection (re ranking)

- Goal. From the retrieved candidate list, select the single best match for the original (German) medication string.
- Approach. We call a second LLM with a strict system prompt that frames the task as a classification over a fixed candidate list. The model is required to output only the index of the best matching list entry, selected on the basis of:

- Active ingredient match (highest priority).
- Strength match (if present in the original name; otherwise avoid candidates that introduce strength). Manufacturer match (only if present in the original name; otherwise avoid introducing it).
- Package size match (only if present; otherwise avoid introducing it).

Tie breakers and abstention policy.

- If there is no candidate that satisfies ingredient and strength constraints, the model prefers entries without conflicting or additional constraints (e.g., selects an ingredient only concept rather than a mismatched strength).
- If no plausible match exists, the model is instructed to return a special index value indicating “no suitable match.”

#### 2.2.2. Implementation details

- We issue up to four queries per record: the original German string and up to three English synonyms. Each query retrieves the top 20 results from the index; candidates are then deduplicated by concept identifier.
- The selection LLM is constrained to output a single integer (the chosen index) or -1. post validation checks: if the emitted index is out of range, the record is flagged for review.

### 2.3. Graph visualization

Neo4J [11] is used to integrate the resulting cross-mapping outputs of both approaches as a property graph by importing cross-mapping tables as relationships between product nodes and RxNorm and RxNorm Extension concept nodes, while preserving provenance attributes (e.g., approach, confidence, review status) on edges. Vocabulary hierarchies and compositional relations (e.g., ingredient-of, has-form, quantified/boxed presentation links, and others) were loaded as additional relationship types, enabling traversal across granularity levels and inspection of one-to-many or many-to-one mapping patterns. Neo4j Bloom provided an interactive exploration layer on top of this graph, allowing users to search by term or identifier, expand neighborhoods along selected relationship types, and visually validate mappings by contextualizing each Swiss product within its connected RxNorm hierarchy and related concepts. Import runs via WinSCP within a MobaXterm network. Access is deeply integrated into the Inselspital’s infrastructure, see overall approach in Figure 2.

**Figure 2.**
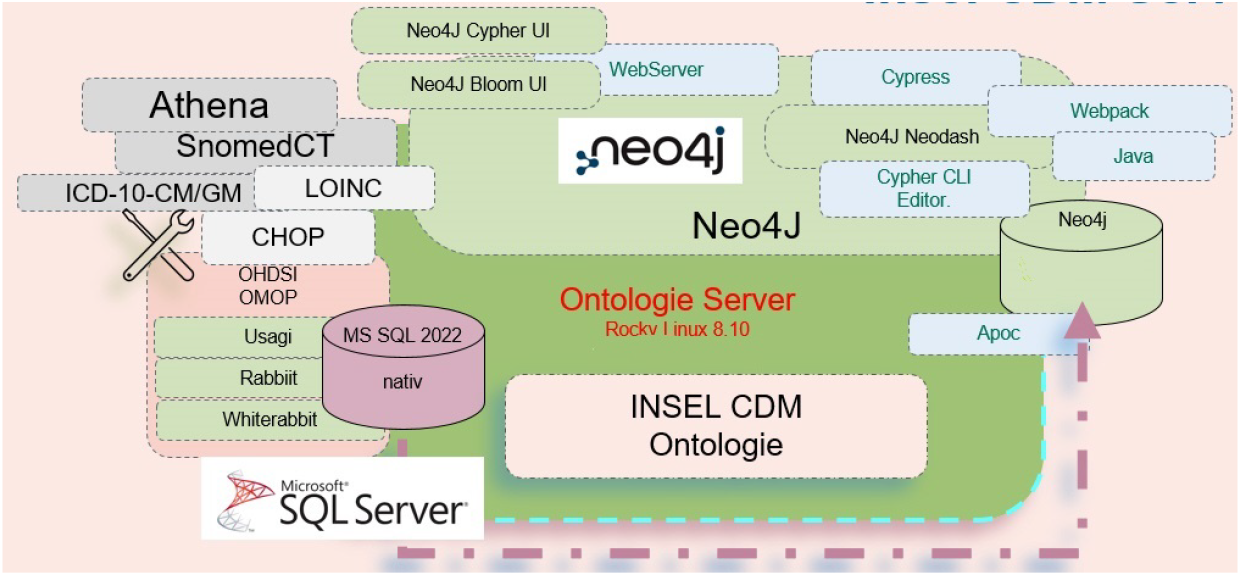
Neo4J Inselspital integration

## 3. Results

### 3.1. Mapping to classes

Across the 151 Swiss medication products not equally mappable on code level, the LLM workflow mapped predominantly to branded-level classes (Branded Drug / Branded Drug Form / Branded Drug Comp / Branded Drug Box / Quant Branded Drug), accounting for 121/151 (80.1 percent) of assignments, whereas the manual Usagi workflow more frequently produced clinical drug–level classes (Clinical Drug / Clinical Drug Form / Clinical Drug Comp / Quant Clinical Drug), totaling 87/151 (57.6 percent). The single most frequent LLM class was Branded Drug Form (71/151, 47.0 percent), followed by Branded Drug Comp (22/151, 14.6 percent) and Branded Drug (19/151, 12.6 percent). In contrast, manual mappings were split between Clinical Drug Form (48/151, 31.8 percent) and Branded Drug Form (46/151, 30.5 percent), with Clinical Drug also common (26/151, 17.2 percent). Notably, the LLM generated Quant Branded Drug (8/151, 5.3 percent) and Branded Drug Box (1/151, 0.7 percent) assignments, which were not used in the manual workflow, while Quant Clinical Drug appeared more often in manual mappings (7/151, 4.6 percent) than in LLM mappings (1/151, 0.7 percent). Non-product-level assignments were rare in both approaches (Ingredient/Marketed Product combined: LLM 5/151, 3.3 percent; manual 3/151, 2.0 percent), indicating that both pipelines generally mapped beyond ingredient-only representations. All results in Table 1 .

**Table 1.** Class comparison of products not equally mappable on code level, LLM and manual USAGI workflow.

| Class | LLM | USAGI manual |
| --- | --- | --- |
| Branded Drug | 19 | 14 |
| Branded Drug Box | 1 | 0 |
| Branded Drug Comp | 22 | 1 |
| Branded Drug Form | 71 | 46 |
| Clinical Drug | 5 | 26 |
| Clinical Drug Comp | 8 | 0 |
| Clinical Drug Form | 11 | 6 |
| Ingredient | 3 | 48 |
| Marketed Product | 2 | 1 |
| Quant Branded Drug | 8 | 2 |
| Quant Clinical Drug | 1 | 7 |
| All | 151 | 151 |

The products equally mappable on code level (28/179, 15.6 percent of total mappings equal and not equall) were mostly mapped to branded-level classes (26/28).

### 3.2. Semantic Proximity

Semantic proximity between mapped concepts was assessed using the median shortest path length in the knowledge graph; shorter paths indicate closer semantic agreement after mapping. Table 2 reports median path lengths stratified by concept class and target vocabulary (RxNorm, RxNorm Extension, and the subset represented in both) for the LLM and manual/USAGI workflows; NV indicates that no values were available for that stratum. Overall, LLM showed shorter median paths for RxNorm (2.47 vs 2.81), whereas manual/USAGI showed shorter median paths for RxNorm Extension (2.46 vs 2.66); for concepts represented in both vocabularies, the median was identical (2.62 vs 2.62). The largest divergence occurred for Branded Drug (RxNorm), where LLM achieved a markedly shorter median path (1.33) than manual/USAGI (5.00); in RxNorm Extension the corresponding medians were 2.50 (LLM) and 2.62 (manual/USAGI), and in the Both subset 2.32 (LLM) versus 2.79 (manual/USAGI). Branded Drug Form remained consistently close across workflows, with LLM shorter in RxNorm (2.10 vs 2.50), but manual/USAGI shorter in RxNorm Extension (2.08 vs 2.54) and in the Both subset (2.15 vs 2.48). For Clinical Drug, LLM was closer in RxNorm (3.00 vs 3.64), while manual/USAGI was closer in RxNorm Extension (3.07 vs 4.00) and marginally closer in the Both subset (3.31 vs 3.40).

**Table 2.** Path length comparison, products not equally mappable on code level, Rx Norm Rx Norm Extension, LLM and manual USAGI workflow: median shortest path length.

| Classes | Rx Norm |  | Rx Norm Extension |  | Both |  |
| --- | --- | --- | --- | --- | --- | --- |
|  | LLM | manual USAGI | LLM | manual USAGI | LLM | manual USAGI |
| Branded Drug | 1.33 | 5.00 | 2.50 | 2.62 | 2.32 | 2.79 |
| Branded Drug Box | NV | NV | 4.00 | NV | 4.00 | NV |
| Branded Drug Comp | 3.25 | NV | 3.06 | 2.00 | 3.09 | 2.00 |
| Branded Drug Form | 2.10 | 2.50 | 2.54 | 2.08 | 2.48 | 2.15 |
| Clinical Drug | 3.00 | 3.64 | 4.00 | 3.07 | 3.40 | 3.31 |
| Clinical Drug Comp | 2.80 | 3.00 | 2.00 | 3.50 | 2.50 | 3.17 |
| Clinical Drug Form | 3.00 | 2.42 | 2.43 | 2.50 | 2.64 | 2.44 |
| Ingredient | 2.00 | 3.00 | NV | NV | 2.00 | 3.00 |
| Marketed Product | NV | NV | 2.50 | 3.00 | 2.50 | 3.00 |
| Quant Branded Drug | NV | NV | 2.75 | NV | 2.75 | NV |
| Quant Clinical Drug | NV | 3.43 | 4.00 | NV | 4.00 | 3.43 |
| All | 2.47 | 2.81 | 2.66 | 2.46 | 2.62 | 2.62 |

Combination product classes showed heterogeneous patterns and more frequent NV values: Clinical Drug Comp favored LLM in RxNorm Extension (2.00 vs 3.50) and in Both (2.50 vs 3.17), whereas Branded Drug Comp had no manual/USAGI values in RxNorm (NV) but favored manual/USAGI in RxNorm Extension and Both (2.00 vs 3.06; 2.00 vs 3.09). Ingredient mappings were available in RxNorm and Both (LLM 2.00 vs manual/USAGI 3.00) but were NV in RxNorm Extension for both workflows. Several packaging/quantification-related classes (e.g., Branded Drug Box, Quant Branded Drug, Quant Clinical Drug) contained NV values. Because of small numbers (179 mappings in total, with 151 mappings having a path length of > 0), we only report median values (Table 2)

### 3.3. Graph visualization

Relationships are presented separately for LLM-based mapping and USAGI mapping. Concept classes can be highlighted. The path length between mapped concepts can be visualized separately for LLM or USAGI workflow enabling a visual comparison. A shorter path corresponds to a better match than a longer path. For exploratory analysis, we compared the distribution of path lengths across mapping approaches to assess differences in semantic proximity. The knowledge graphs contain all relationship types loaded in Neo4J database: MAPS_TO, MAPS_TO_(VIA ATC), IS_A, RXNORM_ING_OF, BRAND_NAME_OF, FORM_OF, RXNORM_INVERSE_IS_A, PRECISE_ING_OF, CONSISTS_OF, HAS_DOSE_FORM_GROUP,

TRADENAME_OF, RXNORM_DOSE_FORM_OF. Examples illustrating both mapping approaches, including cases involving combination products (multi-ingredient formulations) and knowledge graph representations incorporating ATC codes (Nodes: product = orange, Rx Norm = blue, Rx Norm Extension = red; Relationships: red = LLM mapping, blue manual USAGI mapping; Classes: Ingredient, Clinical Drug, Branded Drug), see Figures 3 4 5 6

**Figure 3.**
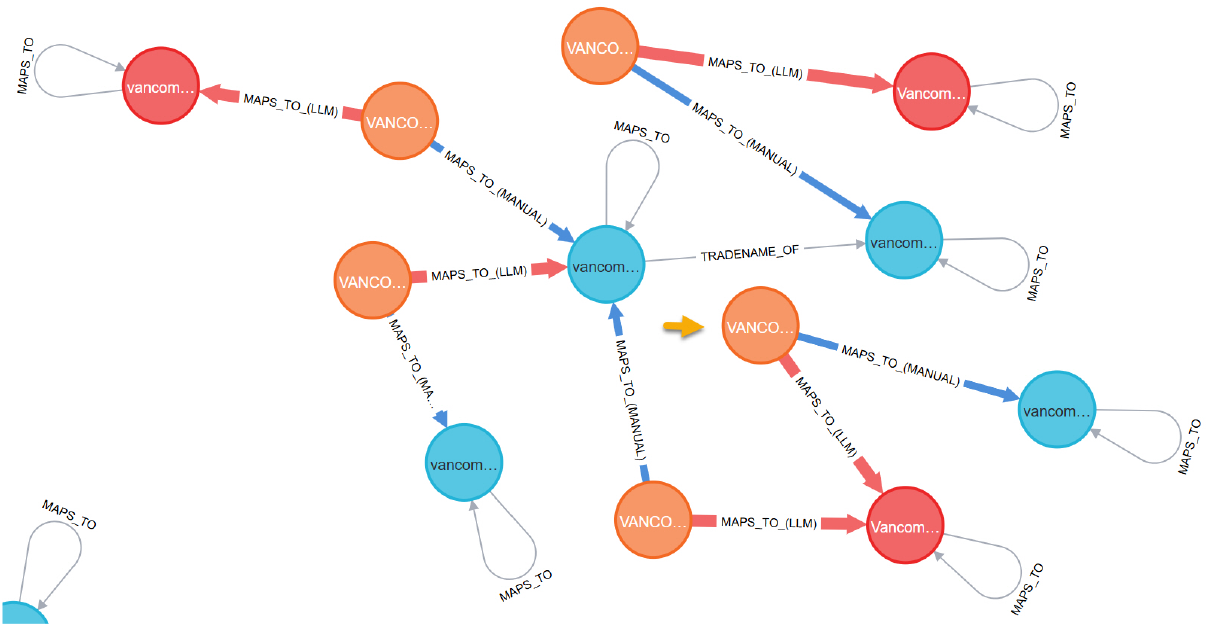
Vancocin 1500 mg infusion; both LLM and manual USAGI mapping

**Figure 4.**
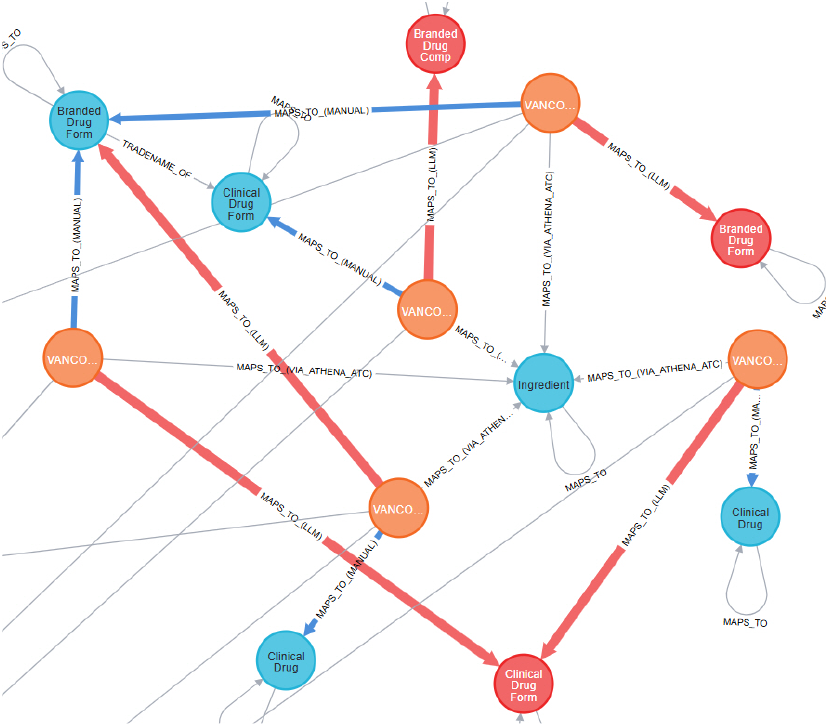
Vancocin 1500 mg infusion, both LLM and manual USAGI mapping and concept classes

**Figure 5.**
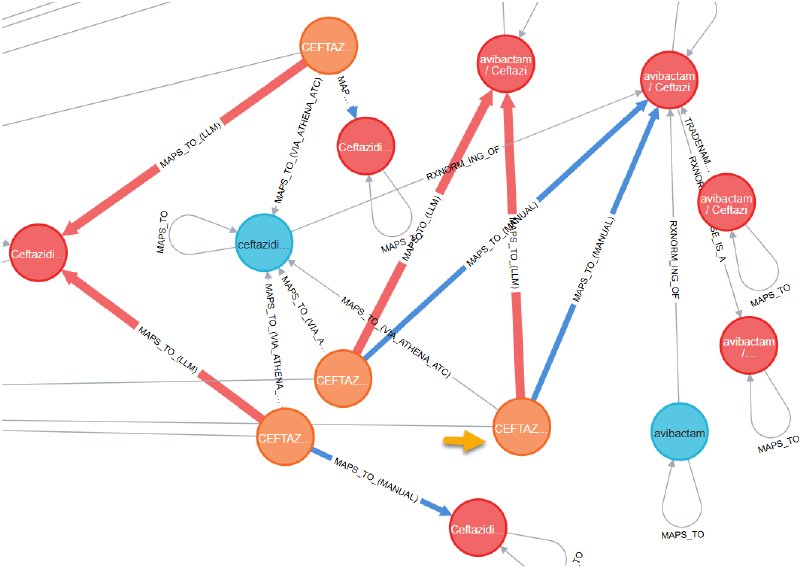
Caeftazidim Avibactam combined formulation

**Figure 6.**
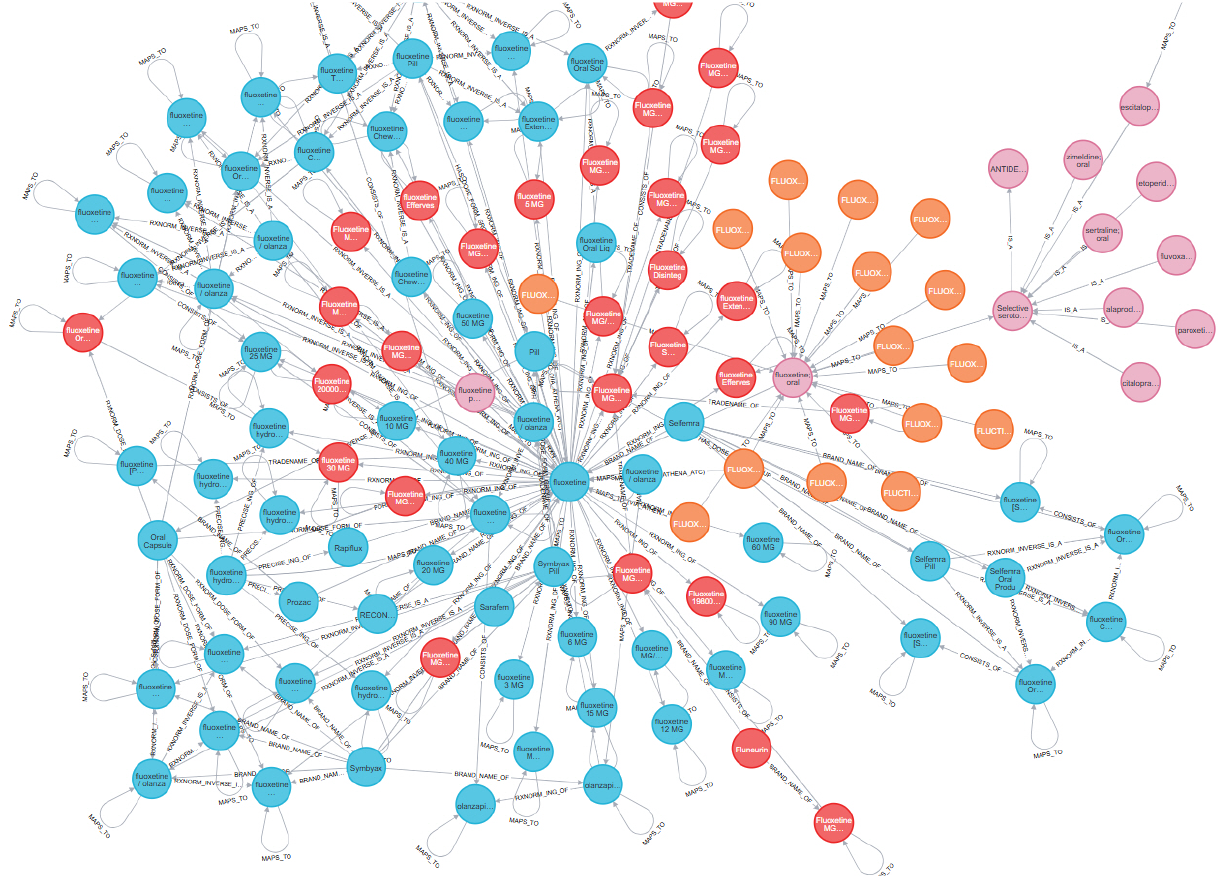
Fluoxetine Knowledge Graph with ATC and all relationsships

## 4. Discussion

Our results confirm that exact product-level normalization of Swiss medication names remains difficult even when mapping to RxNorm and RxNorm Extension. The limited set of fully concordant mappings indicates persistent ambiguity and incomplete target coverage, while the discordant subset shows that many differences reflect selection of different hierarchical levels rather than purely random errors. In the 151 discordant products, the LLM workflow mapped predominantly to branded-level classes (121/151, 80.1 percent), whereas the manual Usagi workflow more often mapped to clinical drug–level classes (87/151, 57.6 percent). The LLM’s frequent assignment to Branded Drug Form suggests a tendency to retain brand-anchored representations, while the manual approach favored Clinical Drug Form and Clinical Drug, consistent with expert choices that prioritize clinically meaningful, less brand-dependent concepts when source strings are under-specified. The appearance of Quant Branded Drug and Branded Drug Box only in the LLM output further points to occasional over-specification toward package-oriented concepts when packaging cues are present in the free text. We introduced a graph-based measure of semantic proximity. Using the median shortest path length between mapped concepts as a proxy for conceptual agreement, we observed a vocabulary-dependent pattern in semantic proximity.

In RxNorm, the LLM workflow achieved the shortest overall median path length (2.47) compared with manual/USAGI mapping (2.81), suggesting that LLM-derived mappings more often connected local product concepts to semantically nearer RxNorm concepts in the integrated knowledge graph. In contrast, for RxNorm Extension, manual/USAGI mapping yielded the shortest overall median (2.46) relative to LLM (2.66), indicating that the advantage of the LLM approach did not uniformly extend to the extension namespace.

The strongest workflow divergence was observed for Branded Drug mappings in RxNorm, where LLM mappings yielded substantially shorter paths (median 1.33) than manual/USAGI mappings (5.00). This is consistent with branded products being particularly sensitive to differences in how trade names, marketed forms, and packaging-related representations are resolved, and it highlights that non-identical code-level mappings can differ markedly in semantic distance. By contrast, Branded Drug Form showed comparatively stable proximity across approaches, with short medians overall; however, the direction of the difference depended on the target vocabulary (RxNorm: LLM 2.10 vs manual/USAGI 2.50; RxNorm Extension: LLM 2.54 vs manual/USAGI 2.08). For Clinical Drug, LLM showed a proximity advantage in RxNorm (3.00 vs 3.64), whereas manual/USAGI performed better in RxNorm Extension (3.07 vs 4.00), indicating that workflow performance and semantic proximity are contingent on both concept class and the target vocabulary.

Combination products (Comp classes) remained a principal challenge: proximity varied by workflow and target vocabulary, and missing strata (NV; no values available) were more frequent. This aligns with the expectation that multi-ingredient formulations are a major driver of non-identical code-level mappings and heterogeneous coverage across RxNorm and its extension.

The outlined discrepancies are plausibly driven by (i) heterogeneous Swiss naming conventions that mix ingredient, strength, form, pack, and manufacturer information in variable ways, and (ii) incomplete RxNorm and RxNorm Extension coverage for Swiss-marketed products, brands, and presentations. When an exact Swiss product is missing, both approaches must approximate by mapping to a less specific concept, often moving upward to clinical drug or ingredient-level entities. In this setting, manual USAGI mapping tends to be more exact because expert adjudication can deliberately avoid introducing unsupported attributes (e.g., strength, route, or pack size) and can select the lowest defensible level given the evidence in the source string. The LLM approach can substantially reduce time spent on translation, normalization, and candidate triage, but it requires careful configuration (prompts, constraints, abstention rules, and post-validation) to prevent overly specific or hallucinated assignments and to align outputs with the intended analytical granularity.

Despite these limitations, RxNorm-based standardization remains advantageous even when only ingredient- or clinical drug–level mapping is achievable. Compared with ATC, RxNorm representations better support interoperability for observational research because they can carry strength and dose-form semantics and, crucially, enable reuse of established crosswalks and roll-ups available in the OMOP CDM or other local vocabulary ecosystem like the Swiss Personalized Health Network (SPHN) . [1] [12] Therefore, an ingredient-level RxNorm mapping can still function as a robust pivot for harmonization and downstream analytics, whereas ATC mappings may remain ambiguous for exposure modeling. At the same time, the observed Swiss gaps reinforce the need for a more comprehensive European (and Swiss) extension to RxNorm that incorporates market-specific products and presentations while remaining OMOP CDM - compatible. Finally, the Neo4j-based graph representation materially improves validation by allowing rapid identification of systematic error modes (e.g., strength mismatches, many-to-one collapses) and by making hierarchical-level gaps visible, which supports targeted remediation and more transparent quality assurance than record-by-record review alone. Graph-based visualization in Neo4j adds substantial value by making provenance, ambiguity, and hierarchical mapping gaps transparent, thereby accelerating validation. In addition, graph visualization supports semantic proximity analyses by enabling inspection of shortest-path connections between mapped concepts. By stratifying proximity patterns by concept class and mapping approach, the graph view helps localize where mismatches cluster and where target vocabulary coverage is insufficient. Finally, interactive traversal of relationships (e.g., ingredient, dose form, marketed form, and packaging links) provides an audit trail that improves interpretability of “near-miss” mappings and informs whether remediation should prioritize mapping rules, source normalization, or vocabulary extension. This representation enabled a knowledge graph for not only the study’s mapped products but also integrated classifications beyond the objectives of this study, i.e. ATC.

## 5. Conclusion

RxNorm and the RxNorm Extension provide a strong interoperability anchor for medication standardization because they support normalized drug identifiers and enable reuse of established OMOP CDM vocabulary relationships and crosswalks beyond what ATC can offer. However, the observed coverage gaps for Swiss and broader European products underscore the need for a dedicated European/Swiss extension that systematically represents locally marketed drugs, presentations, and packaging conventions. The LLM-based workflow demonstrated high operational efficiency and plausible mapping quality, but it requires domain-specific constraints, calibration, and quality controls to avoid over-specific or inconsistent hierarchical assignments. Graph-based visualization in Neo4j adds substantial value by accelerating validation, supporting semantic proximity analyses and by providing an audit trail that improves interpretation of “near-miss” mappings. Serving as a knowledge graph it helps to understand dependencies and gaps beyond the objectives of this study. Next, we will scale the approach to the full catalog using a class-by-class mapping strategy, beginning at the highest hierarchical level and progressively refining mappings toward more specific concept classes where sufficient source evidence and target coverage exist.

## Acknowledgments

We want to thank IT-Logix AG, Bern, Switzerland for support and Marcel Affolter for all the patience he needed when setting up the technical infrastructure.

## Data availability statement

All data supporting the findings of this study are available within the paper and its Supplementary Information.

## Declaration on Generative AI

During the preparation of this work, the author(s) used GPT-5.1 and DeepL for style, grammar, and spelling checks. After using these tools, the authors reviewed the complete content and edited the content as needed for corrections. The authors take full responsibility for the publication’s content.

## Notes

### Competing Interest Statement

The authors have declared no competing interest.

### Funding Statement

This study did not receive any funding

